# Randomised, double-blind, placebo-controlled pilot clinical trial of jackfruit seed extract formulation (JaSE) for allergic rhinitis treatment

**DOI:** 10.1101/2025.08.24.25334210

**Authors:** Gauri Purohit, Pranali Dayalkar, Rohit Bhat, Jyoti Iyer, Sunil Nadkarni, Omkar Shahapurkar

## Abstract

Allergic rhinitis (AR), a chronic inflammatory disease associated with comorbidities, adversely affects life’s quality, productivity and increases financial healthcare burden. Herbal medicines are emerging for AR treatment. The presence of two immunomodulatory lectins (jacalin and artinM) in jackfruit (*Artocarpus heterophyllus*) seeds suggests a probable therapeutic potential of jackfruit seed extract in AR management. A double-blind, placebo-controlled pilot trial of jackfruit seed extract formulation (JaSE) was conducted in 60 AR patients to assess its AR treatment efficacy by evaluating changes in AR symptoms (score-based) over 15 days of treatment. The study was approved by the Institutional Ethics Review Committee and was registered with CTRI (CTRI/2023/08/056311). Out of 60 patients enrolled, 53 completed the trial. No adverse events were reported throughout the study. Post-treatment, both the JaSE and placebo (saline) groups demonstrated statistically significant reduction in AR symptoms. The potential of JaSE in relieving AR symptoms was observed, though non-significant as compared to placebo. Absence of statistically significant difference between two treatment groups may be attributed to the placebo effect and small sample size. Expanding sample size and conducting adequately powered clinical trials in future will be essential to accurately assess the efficacy of the novel candidate JaSE in AR management.

## Introduction

Allergic rhinitis (AR) is a chronic inflammatory disease caused by an IgE-mediated immunological response of the nasal mucosa. AR, the most common form of allergic disorders, is a global health problem, as 1.4% to 39.7% of the Western population and 20% to 30% of the Indian population suffer from AR.^[1]^ The incidence of AR has greatly increased over the last decade, leading to a social and economic burden. In India, it is the most common reason for visits to health practitioners.^[2]^ AR is characterised by nasal obstruction, itching (nasal, conjunctival, and pharyngeal), watery rhinorrhoea (anterior and posterior), lacrimation, and sneezing. It accompanies other comorbid conditions such as asthma, sinusitis, atopic dermatitis, and nasal polyps.^[3]^ Reports suggest that around 15% of AR cases develop asthma. ^[1]^ Although AR is not life-threatening, due to its associated comorbidities, impaired quality of life, reduced productivity, and treatment cost to the patient, AR is a cause of concern.^[4]^

Available treatments for AR include antihistamines, corticosteroids, chromones, decongestants, anticholinergics, anti-leukotrienes, and immunotherapy.^[1]^ AR medicines can cause many adverse effects such as drowsiness, headache, dry mouth and nose, pyrexia, cough, epistaxis, sneezing, nasal burning and irritation, eye redness, and others. Despite advancements in AR therapy over the years, a fast and full recovery of all patients without any side effects post-treatment has not been achieved yet.^[5]^ Due to the associated side effects of available AR treatments, herbal medicines are emerging as an alternative. A variety of herbal formulations are available for treating AR.^[6]^ However, jackfruit seed extract has not been considered for AR treatment. Jackfruit seeds, which are rich in nutrients, are consumed all over the world and have been proven to have antioxidant, prebiotic, and anticancer potential^.[7]^ Jackfruit seed contains two immunomodulatory lectins (jacalin and artinM).^[8–15]^ Studies on the PBS-soluble material of jackfruit seeds indicated that jacalin comprised more than 50% of PBS-soluble fraction.^[8,9]^ Jacalin binds to SIgA, a key effector molecule in mucosal immune response,^[10]^ induces lymphocyte proliferation in human PBMC and CD4+ T cells,^[11]^ and interacts with CD45 on T cells, mediating T cell activation, and Th1/Th2 cytokine secretion.^[12]^ It also induces the secretion of interferon-γ, IL-2, and IL-6.^[13]^ ArtinM induces neutrophil activation and migration by haptotaxis,^[14–16]^ induces macrophages to produce IL-12 p40,^[17]^ plays a role in mast cell degranulation^[18]^ and interleukin-12 production in a toll-like receptor 2-dependent mechanism.^[19]^ It induced switches from Th2 to Th1 cell-mediated immunity against Leishmania’s major antigens.^[17]^ ArtinM is emerging as a novel immunotherapeutic molecule as it modulates effector function and provides resistance to some intracellular pathogens.^[15]^ The cellular and humoral immune response of jackfruit lectins, jacalin, and artinM against microbial infection has been evaluated *in-vitro* and *in-vivo.*^[17,20–28]^ The synergistic effect of jacalin and artinM is also reported.^[29]^ The presence of these lectins in the jackfruit seed extract suggests probable therapeutic potential in the treatment of AR.

AR involves the impaired response of CD4+T cells to apoptotic stimuli and Th2 polarisation.^[30]^ Recent studies on the effect of allergen-specific immunotherapy on AR in a mice model revealed improved Th1/Th2 cell balance post-treatment and an increased number of CD4+ T cells in peripheral blood.^[31]^ Therefore, the presence of immunomodulatory and antimicrobial lectins jacalin and artinM in jackfruit seed extract suggests its possible role in treating rhinitis. Ayurveda also reports the property of jackfruit seed as an expectorant of mucus, promoting secretion and/or expulsion of mucus.^[32]^ Ayurvedic literature suggests the efficacy and safety of nasal ingestion of jackfruit seed extract.^[32–34]^ Locals also promote the consumption of jackfruit seeds due to the wide range of health benefits of jackfruit seeds.

Therefore, this study aimed to examine the potential of jackfruit seed extract formulation (JaSE) as nasal drops to remedy AR and other ailments related to nasal congestion. Changes in rhinitis and other nasal congestion symptoms post administration of JaSE through the nasal route were monitored during the study. The study was conducted in accordance with applicable Good Clinical Practice and was approved by the Institutional Ethics Review Committee (CTRI/2023/08/056311).

## Materials and Methods

### Study population

For the pilot study, 60 AR patients (30 drug + 30 placebo) were recruited.

### Inclusion criteria

Patients suffering from AR, aged between 18 years and 80 years, who were willing to participate in the study were included.

### Exclusion criteria

Patients with hypersensitivity to jackfruit or jackfruit seed products were excluded from the study. Patients having co-morbidities, pregnant and breastfeeding women, were also excluded. Participants with chronic sinusitis, severe asthma, and on antihistamines, corticosteroid nasal drops, or any other AR treatment medication were not included in the study. At the time of enrolment, patients who were participating in another clinical trial were also excluded from the clinical trial.

### Withdrawal criteria

Withdrawal of informed consent, evidence of the development of other complications, development of exclusion criteria during the course of the study, and protocol non-compliance led to the withdrawal of the participant from the study.

Patients were advised to report immediately if they observed any adverse effects of the drug or an increase in the severity of their AR symptoms during the study. In the event of any adverse reaction, the patient was to be withdrawn from the study.

The investigator ensured that the patient received full and adequate oral and written information about the nature, purpose, possible risks, and benefits of the study. Informed consent was obtained from all participants before the initiation of the study.

### Study design

The study was a randomised, double-blind, placebo-controlled pilot trial conducted between August 2023 and December 2024, at the B. K. L. Walawalkar Hospital, Diagnostic & Research Centre, Ratnagiri, Maharashtra, India.

At the first visit, patients who met the selection criteria were enrolled in the study (Day 1 of the treatment). The investigator examined the eligible patients and recorded their observations. The participants were asked to fill out a questionnaire to access and record their pre-treatment symptoms. The questionnaire was available in the local language. Additionally, before beginning the treatment, blood samples were taken from the participants to access their ESR and CRP. The patients were advised to administer 3 nasal drops per nostril thrice a day, for 15 days. After completing all required procedures, a concealed drop bottle containing either a drug or a placebo was provided to the patients. As the study was double-blind, the investigator and patient were unaware of the content of the drop bottle allocated to the patients. The allocation schedule of the drug/placebo was generated by an independent statistician. A code was given by the statistician to each drop bottle, and a record of the code of the bottle given to the patient was maintained.

On the 2^nd^ visit (Day 15 of the treatment), patients were evaluated for post-treatment symptoms, and the blood sample was also collected for ESR and CRP estimation. Assessment of post-treatment symptoms was based on feedback from patients and clinical investigations conducted by the investigator. On the 3rd visit (Day 30 of the treatment), patients were evaluated for general well-being and the emergence of adverse effects, if any.

The questionnaire given to the patients on the 1st and 2nd visits consisted of questions related to symptoms. Each symptom was rated on a scale from 0 to 4, with higher scores reflecting increased severity. Symptoms considered for evaluation of the treatment were runny nose, blocked nose, high temperature, cough, sore throat, watery eyes, and chest congestion. The nasal obstruction score was also calculated based on rating the severity from 0 to 4 for the following symptoms: Nasal congestion or stiffness, Nasal blockage or obstruction, Trouble breathing through the nose, Trouble sleeping, and Unable to get enough air through the nose during exercise or exertion. The ratings of these symptoms were summed up and multiplied by 5 to calculate nasal obstruction score for the participant. Based on the nasal obstruction score, the AR severity was classified as Mild (5-25), Moderate (30-50), Severe (55-75), and Extreme (80-100).^[35]^

The clinical efficacy of the treatment was assessed by comparing AR symptom ratings, nasal obstruction scores, and clinical evaluations between the 1^st^ and 15^th^ day of treatment. Additionally, ESR and CRP levels were also evaluated.

### Study drug

The study drug was a saline extract of jackfruit seeds (JaSE). Jacalin was considered an active ingredient of the formulation. Hence, protein concentration and SDS-PAGE profile with marker bands of jacalin were considered for standardisation of the JaSE formulation. The standard operating procedures were maintained during the preparation of JaSE. Sterile saline solution (0.9% w/v) was used as a placebo.

### Data analysis

Pre-and post-treatment data were summarised using central tendencies (mean, median), range, standard deviation, and 95% confidence interval (95% CI). Statistical tests were carried out to compare the two treatment groups based on their distribution (normality) and the type & level of measurement of the variable under consideration using parametric and non-parametric tests (like Student’s t-test, Chi-squared test, Wilcoxon Signed-Rank test). The result of statistical analysis was considered significant at P < 0.05 (two-sided). Standard statistical software programs such as IBM SPSS Version 23 were used.

## Results

An overview of the study is presented in the CONSORT flow chart (Figure 1). 60 patients having AR symptoms were enrolled in this study. Out of these participants, 53 (88.33%) patients attended both the 2^nd^ and 3^rd^ follow-up visits and hence, were considered as trial-completed participants. The unblinding of the study revealed that out of a total of 53 participants completing the trial, the JaSE (drug) group had 26 subjects, while the saline (placebo) group had 27 subjects. Seven participants (four of whom received JaSE and three of whom received saline) were considered dropouts during the study as they could not adhere to the study protocol. None of the dropouts was due to drug-related adverse events. There were no medical complications or deaths in the participants during the study.

**Figure 1:**
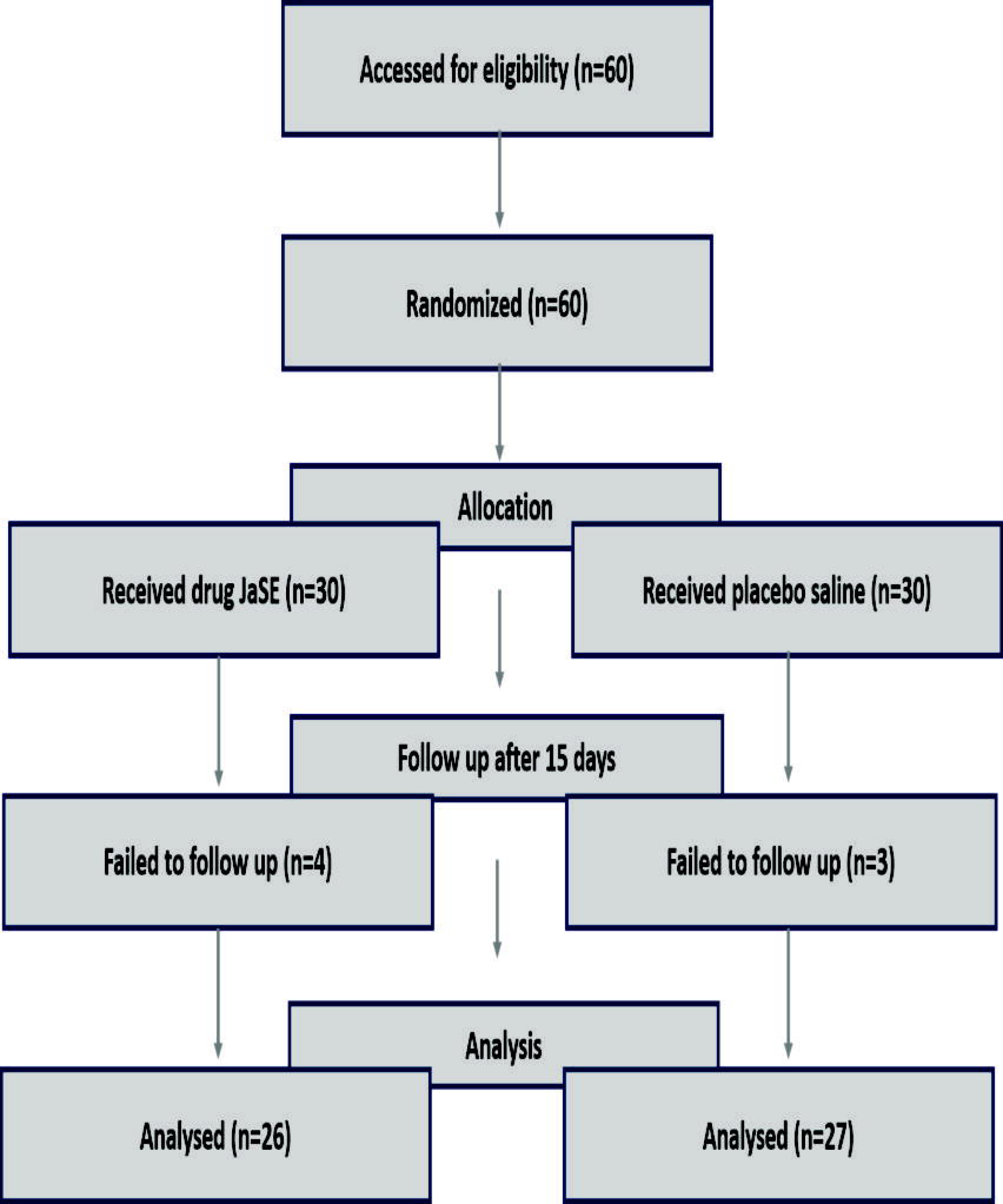
CONSORT flow chart

The baseline characteristics (gender and age) of the trial-completed participants among the placebo and drug groups had no significant difference (Table 1). In the drug group, female gender was dominant, suggesting a slight gender imbalance; however, the distribution of males and females in the placebo group did not show a significant difference. The gender difference between the two treatment groups is noticeable but not extreme. The treatment group did not have any statistically significant associations with gender. The minimum age of the participant was 21 years, and the maximum age was 73 years. The majority of participants were within the 25 to 50 age range, with a noticeable peak around 40 years, and a few older individuals (above 60). The mean age of participants in the drug group was 41.96, while in the placebo group, it was 37.40, indicating a similar age distribution in the two treatment groups.

**Table 1:** Baseline characteristics of treatment groups.

| Indicator |  | JaSE<br>(Drug) | Saline<br>(Placebo) | P-value |
| --- | --- | --- | --- | --- |
| Gender, n (%) | Male | 8 | 13 | 0.311 |
|  | Female | 18 | 14 |  |
| Age (y) | Mean (SD) | 41.962<br>(14.11) | 37.40 (10.25) | 0.306 |
|  | Median (Min, Max) | 42.500 (22, 73) | 36 (21, 62) |  |
| AR symptoms |  |  |  |  |
| Nasal Congestion | Median (Min, Max) | 4 (0, 4) | 4 (0, 4) | 0.179 |
| Nasal Blockage | Median (Min, Max) | 4 (0, 4) | 4 (0, 4) | 0.411 |
| Trouble breathing | Median (Min, Max) | 2 (0, 4) | 2 (0, 4) | 0.985 |
| Trouble sleeping | Median (Min, Max) | 0 (0, 4) | 0 (0, 4) | 0.613 |
| Unable to get enough air | Median (Min, Max) | 0 (0, 4) | 0 (0, 4) | 0.912 |
| Nasal Obstruction score | Median (Min, Max) | 40 (0, 90) | 55 (20,100) | 0.318 |
| Sneezing attacks | Median (Min, Max) | 4 (0, 4) | 4 (0, 4) | 0.645 |
| Headache | Median (Min, Max) | 1.5 (0, 4) | 4 (0, 4) | 0.559 |
| Heaviness of head | Median (Min, Max) | 1.5 (0, 4) | 0 (0, 4) | 0.984 |
| Throat irritation | Median (Min, Max) | 0.5 (0, 4) | 2 (0, 4) | 0.498 |
| Cough | Median (Min, Max) | 0 (0, 4) | 2 (0, 4) | 0.326 |
| Dyspnoea | Median (Min, Max) | 0 (0, 4) | 0 (0, 4) | 0.627 |

The baseline nasal obstruction score and AR symptoms rated from 0 to 4 among the two groups were comparable (Table 1). Their difference was statistically non-significant (Figure 2a and Figure 2b). Based on the nasal obstruction score, it was observed that 12 (drug 7, placebo 5), 15 (drug 7, placebo 8), 9 (drug 3, placebo 6), and 17 (drug 9, placebo 8) participants had mild, moderate, severe, and extreme AR symptoms respectively before starting the treatment.

**Figure 2:**
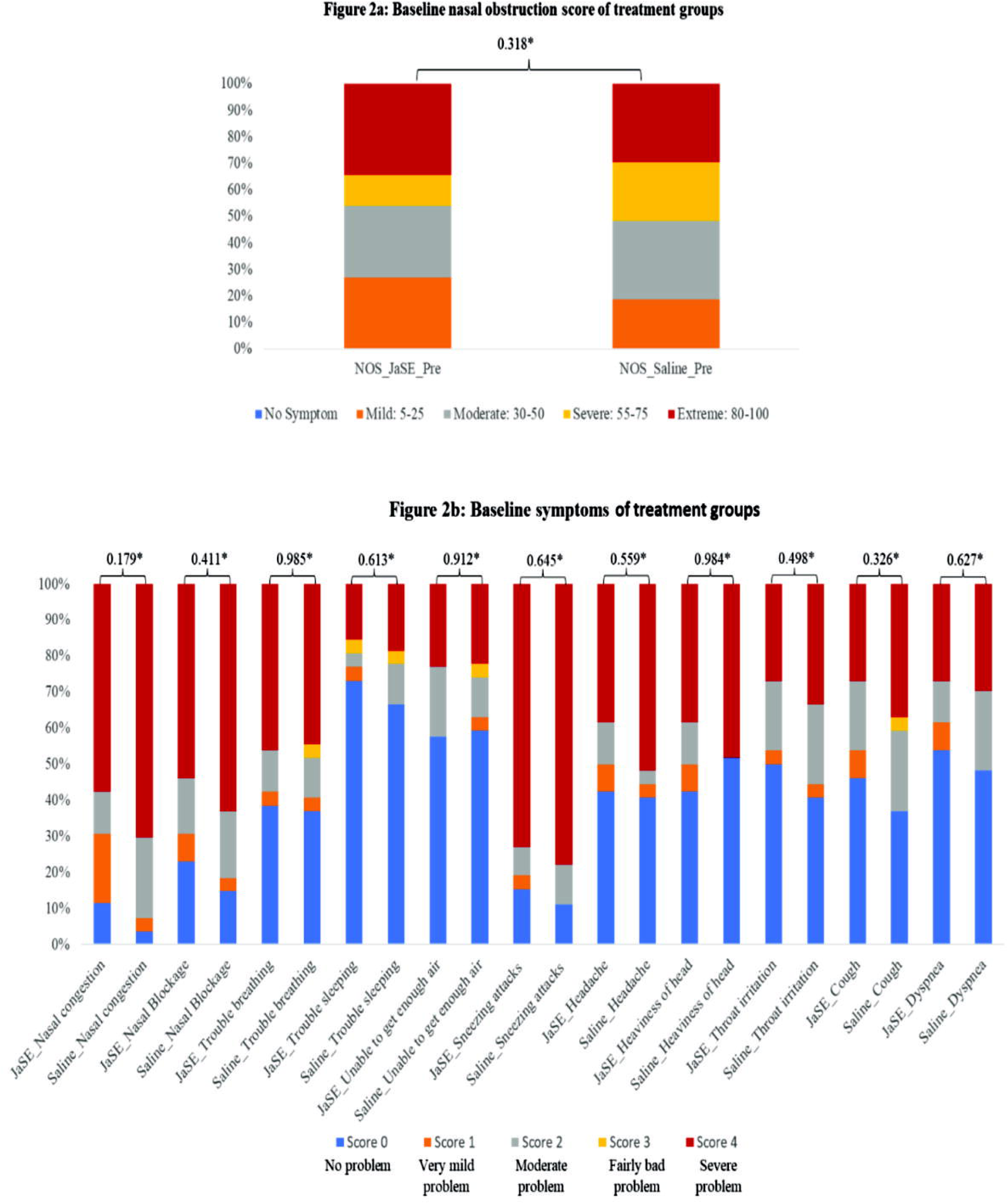
Figure 2a: Baseline nasal obstruction score of treatment groups Figure 2b: Baseline symptoms of treatment groups

For efficacy assessment, changes in the symptom scores after 15 days of treatment, rated by participants and evaluated by the investigator, were recorded. Pre-and post-treatment data analysis among the JaSE-treated subjects revealed a statistically significant (p-value < 0.05) improvement in the symptoms (Table 2). Every symptom showed significant improvement post-treatment with JaSE. No subject reported any adverse event of the treatment. Nasal obstruction score analysis of the participants in the JaSE group showed that 0, 7, 7, 3, and 9 participants had no, mild, moderate, severe, and extreme symptoms, respectively, before starting the treatment. However, post-treatment, the number changed to 12, 7, 2, 1, and 4 for no, mild, moderate, severe, and extreme symptoms, respectively (Figure 3a).

**Figure 3:**
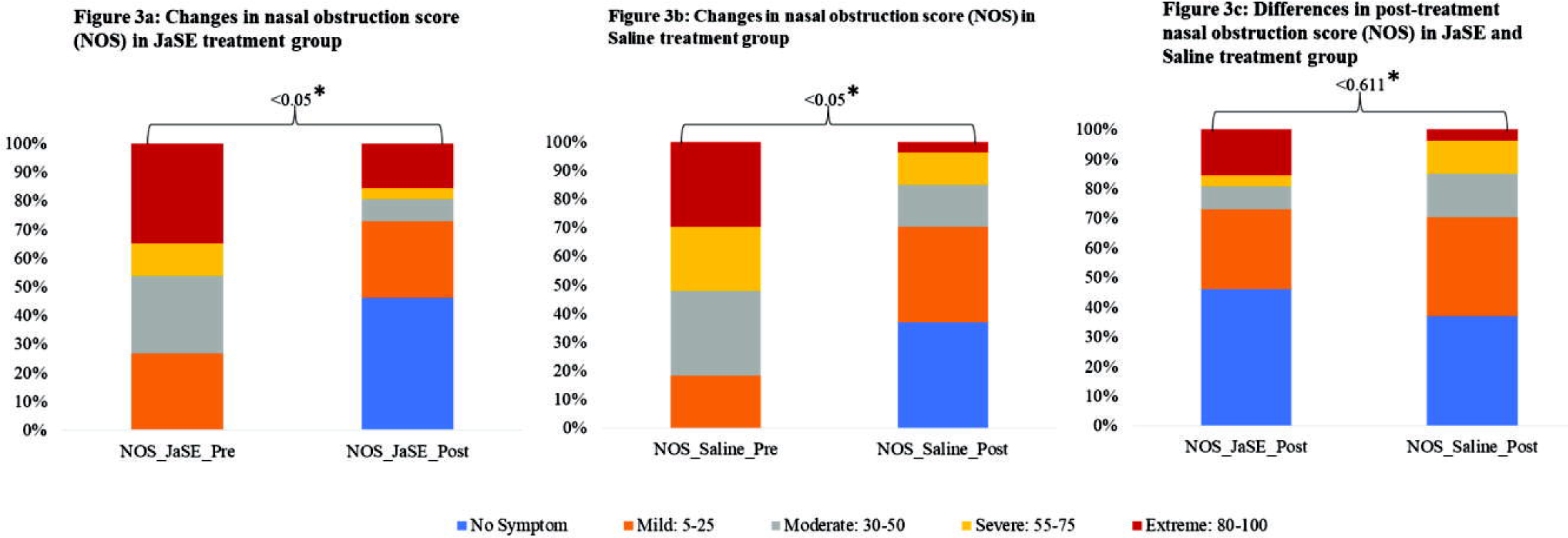
Figure 3a: Changes in nasal obstruction score (NOS) in JaSE treatment group Figure 3b: Changes in nasal obstruction score (NOS) in Saline treatment group Figure 3c: Differences in post-treatment nasal obstruction score (NOS) in JaSE and Saline treatment group

**Table 2:**
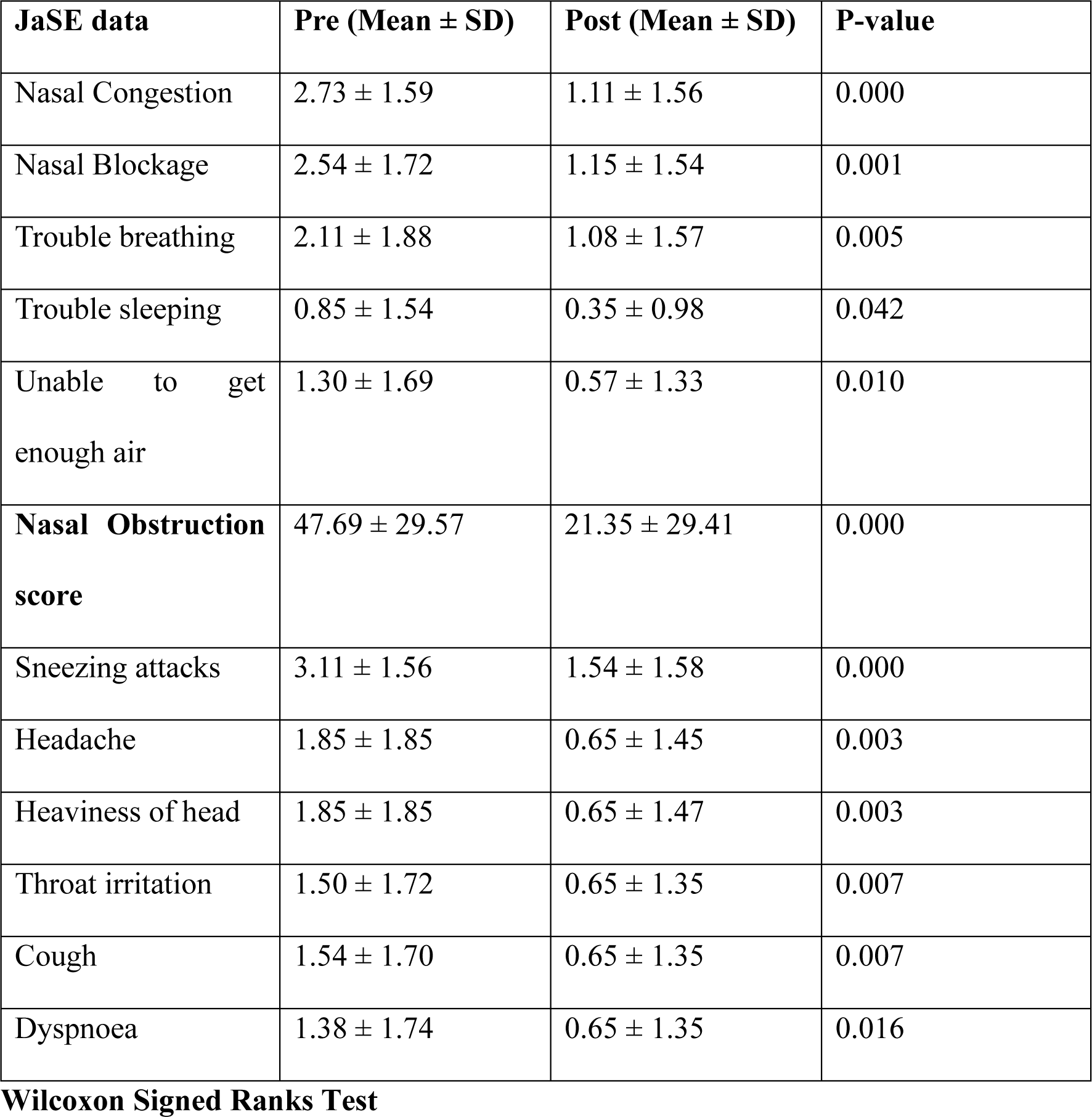
Changes in symptoms in JaSE treatment group.

Analysis of the placebo group revealed that saline treatment also showed statistically significant (p-value < 0.05) improvement in AR symptoms post-treatment (Table 3). No subject reported any side effects of the treatment. Nasal obstruction score analysis of the participants in the saline group showed that 0, 5, 8, 6, and 8 participants had no, mild, moderate, severe, or extreme symptoms, respectively, before starting the treatment. However, post-treatment, the number changed to 10, 9, 4, 3, and 1 for no, mild, moderate, severe, and extreme symptoms, respectively (Figure 3b).

**Table 3:** Changes in symptoms in Saline treatment group.

| **Saline data** | **Pre (Mean ± SD)** | **Post (Mean ± SD)** | **P-value** |
| --- | --- | --- | --- |
| Nasal Congestion | 3.30 ± 1.17 | 1.37 ± 1.60 | 0.000 |
| Nasal Blockage | 2.92 ± 1.54 | 1.18 ± 1.54 | 0.000 |
| Trouble breathing | 2.15 ± 1.85 | 0.56 ± 1.09 | 0.001 |
| Trouble sleeping | 1.07 ± 1.63 | 0.30 ± 0.76 | 0.016 |
| Unable to get enough air | 1.26 ± 1.70 | 0.70 ± 1.26 | 0.027 |
| **Nasal Obstruction score** | 55.74 ± 25.71 | 22.59 ± 26.36 | 0.000 |
| Sneezing attacks | 3.33 ± 1.36 | 1.56 ± 1.65 | 0.000 |
| Headache | 2.18 ± 1.96 | 1.22 ± 1.65 | 0.007 |
| Heaviness of head | 1.92 ± 2.04 | 1.07 ± 1.66 | 0.016 |
| Throat irritation | 1.81 ± 1.75 | 0.81 ± 1.47 | 0.004 |
| Cough | 2.04 ± 1.76 | 1.04 ± 1.22 | 0.002 |
| Dyspnoea | 1.63 ± 1.76 | 0.63 ± 1.18 | 0.004 |

While both the JaSE (drug) and saline (placebo) groups showed symptomatic improvement, the difference in the magnitude of improvement between the two treatment groups was not statistically significant, indicating that the superiority of JaSE over placebo could not be established at this stage (Table 4). A slightly greater number of participants in the JaSE group (12 participants) reported complete symptom resolution compared to the saline group (10 participants) (Figure 3c). This observed improvement, though not significant, may suggest a therapeutic effect of JaSE, which warrants further investigation.

**Table 4:** Comparison of two treatment groups post 15 days of treatment.

| **AR symptoms post-treatment** | **Drug** | **Placebo** | **P-value** |
| --- | --- | --- | --- |

|  | Median<br>(Min,<br>Max) | Post (Mean<br>± SD) | Median<br>(Min,<br>Max) | Post (Mean<br>± SD) |  |
| --- | --- | --- | --- | --- | --- |
| Nasal<br>Congestion | 4 (0, 4) | 1.11 ± 1.56 | 4 (0, 4) | 1.37 ± 1.60 | 0.485 |
| Nasal Blockage | 4 (0, 4) | 1.15 ± 1.54 | 4 (0, 4) | 1.18 ± 1.54 | 0.977 |
| Trouble<br>breathing | 2 (0, 4) | 1.08 ± 1.57 | 2 (0, 4) | 0.55 ± 1.08 | 0.186 |
| Trouble<br>sleeping | 0 (0, 4) | 0.35 ± 0.98 | 0 (0, 4) | 0.30 ± 0.77 | 0.943 |
| Unable to get<br>enough air | 0 (0, 4) | 0.58 ± 1.33 | 0 (0, 4) | 0.70 ± 1.26 | 0.465 |
| **Nasal**<br>**Obstruction**<br>**score** | 7.5 (0, 80) | 21.35±<br>29.41 | 15 (0,<br>100) | 22.59 ±<br>26.36 | 0.611 |
| Sneezing<br>attacks | 4 (0, 4) | 1.54 ± 1.58 | 4 (0, 4) | 1.55 ± 1.65 | 0.985 |
| Headache | 1.5 (0, 4) | 0.65 ± 1.47 | 4 (0, 4) | 1.22 ± 1.65 | 0.080 |
| Heaviness of<br>head | 1.5 (0, 4) | 0.65± 1.47 | 0 (0, 4) | 1.07 ± 1.66 | 0.192 |
| Throat<br>irritation | 0.5 (0, 4) | 0.65 ± 1.35 | 2 (0, 4) | 0.81 ± 1.47 | 0.613 |
| Cough | 0 (0, 4) | 0.65 ± 1.35 | 2 (0, 4) | 1.04 ± 1.22 | 0.077 |
| Dyspnoea | 0 (0, 4) | 0.65 ± 1.35 | 0 (0, 4) | 0.63 ± 1.18 | 0.722 |

Due to the unwillingness of the participants to provide blood samples after treatment, only 28 participants’ pre-and post-treatment data for CRP and ESR could be collected. However, there were insignificant changes in CRP and ESR values, and no correlation could be established.

## Discussion

In the current pilot study, it was observed that JaSE could relieve AR symptoms in the participants. Nasal drops were easy to take and patients did not report any adverse events after administering the JaSE nasal drops, indicating its safety. Significant relief was observed for all symptoms considered during the study after administering JaSE drops. Patients reported that the frequency of occurrence of rhinitis and intensity of its symptoms reduced post-15 days of treatment with JaSE.

However, comparing symptomatic relief in JaSE and saline treatment groups does not prove the superiority of any one treatment group. Still, the JaSE group had a slightly higher number of patients with no symptoms post-treatment than the saline group. The absence of a significant difference between the two groups might be due to the placebo effect^36^ as well as due to saline’s role in relieving rhinitis symptoms.^[37]^ An increase in the number of participants in the study will help in establishing the efficacy of JaSE. Based on differences in nasal obstruction scores between the two treatment groups, it is estimated that a higher sample size (≈ 455 with 95 % confidence and 80 % power) is required for further study. Documentation of changes in the quality of life of patients and a daily log of changes in symptoms was not performed during the current study. This data collection will demonstrate the time-dependent efficacy of JaSE.

A wide range of products are available for the treatment of AR however, there are some side effects associated with them. Hence, an alternative is needed, and JaSE is indicating its potential as an alternative. This clinical trial revealed that JaSE was safe and effective in patients with AR. Optimizing the protocol and JaSE dosing is expected to increase and maintain the efficacy of this novel approach. The lack of significant findings may, in part, be due to the placebo effect and the limited sample size. Expanding the sample size and conducting adequately powered clinical trials in future will be essential to accurately assess the efficacy of JaSE in AR management.

The current formulation, which is derived from Ayurvedic literature and scientific evidence, will assist in relieving rhinitis symptoms without any adverse effects. Biomolecules present in JaSE would help to reduce nasal mucosa inflammation, reduce microbial load, and clear airways. Further work on JaSE is necessary to determine the exact degree of effectiveness. Our opinion is that JaSE treatment has the potential to significantly enhance the quality of life for AR sufferers everywhere.

## Data Availability

All data produced in the present study are available upon reasonable request to the authors

## Acknowledgement

The authors express sincere gratitude to Dr Suvarna Patil, Medical Director for timely support and guidance. Dr Sonam Kotawdekar is acknowledged for her support in the execution of the project. The authors thank Mr Charudatta Joglekar for help in blinding and unblinding the coded drug/placebo bottles used in the study. Miss Tanishka Khedkar and Miss Shruti Taraye are acknowledged for their support in JaSE preparation and conducting this clinical trial.

